# Quantification of droplet and contact transmission risks among elementary school students based on network analyses using video-recorded data

**DOI:** 10.1101/2024.10.25.24316099

**Authors:** Shuta Kikuchi, Keisuke Nakajima, Yasuki Kato, Takeshi Takizawa, Junichi Sugiyama, Taisei Mukai, Yasushi Kakizawa, Setsuya Kurahashi

## Abstract

Elementary schools are environments in which immunologically immature students come into close contact with each other and are susceptible to the spread of infectious diseases. Analyzing the behavior of multiple students has been challenging, and the relationship between infections remains unclear. In this study, we analyzed the relevance between droplet and contact transmission and the behavior of elementary school students using video-recorded data, network analyses, and simulations. The analysis of communication behavior revealed the diverse nature of interactions among students. By calculating the droplet transmission probabilities based on conversation duration, this study quantified the risk of droplet transmission in elementary schools. The analysis of contact behavior introduces a novel approach for constructing contact networks based on contact history. According to this method, items such as desks, shirts, and doors have the potential to be used as fomites for virus transmission. In addition, the reliability of the predictions was demonstrated through micro-simulations. Interestingly, the micro-simulations indicated that the majority of virus copies were transmitted through single items, emphasizing the importance of targeted hygiene measures. This study contributes significantly to the prevention of infectious diseases in elementary schools by providing evidence-based information on transmission pathways and behavior-related risks. Moreover, the insights from this study can guide the development of simulation models for analyzing infection risks in educational settings.

## Introduction

Human respiratory viruses cause significant morbidity, mortality, and economic losses globally each year [1–4]. Occasional pandemics, such as the COVID-19 pandemic and the 2009 H1N1 influenza pandemic, have severely disrupted society and economics. Viruses can be transmitted mainly via two pathways [5, 6]. The first is droplet transmission, which occurs through exposure to droplets produced by coughing, sneezing, and conversations with virus carriers. Based on their size, these droplets can be further categorized into larger droplets and fine aerosols. The second pathway is contact transmission, which occurs when a virus carrier comes into contact with surfaces contaminated with droplet-borne viruses. Contact transmission can be further categorized into direct contact, which involves direct physical contact, and indirect contact, which involves contact with fomites. Droplet and contact transmissions typically spread through social networks [7–10].

Schools are considered an important area for virus transmission because of the close contact between students, teachers, and school staff [11–15]. Furthermore, infected elementary school students can also become a source of infection within their households, and these infections can spread throughout the community [12, 16–18]. Therefore, investigating virus transmission in elementary schools is important from the perspective of preventing outbreaks.

To estimate virus transmission in elementary schools, it is necessary to obtain behavioral data related to droplet and contact transmissions from students. For droplet transmission, data on the duration of conversations between students are required, whereas for contact transmission, data on the items they contact are required. To investigate the relationship between elementary school students and droplet transmission, previous studies collected communication data using questionnaires and wearable sensor devices [11, 19–22]. However, a questionnaire cannot capture details such as the type, time, and frequency of communication in actual behaviors. In addition, wearable sensor devices can only detect proximal contact and cannot determine whether a conversation took place. It is also difficult to obtain information about contacted items and the order and frequency thereof by means of a questionnaire. Research on contact transmission in elementary schools has been limited to methods that detect pathogens present on the surfaces of school items [23]. Therefore, in this study, video recordings of student behavior were employed to obtain detailed behavioral data related to droplet and contact transmission.

In terms of communication behavior, networks among students and the duration of conversations were investigated. Droplet transmission probability was calculated based on the cumulative conversation duration for each pair of students. For contact behavior, the contact items and their frequencies were investigated. In addition, we developed a novel analysis method that constructs networks based on contact history. Fomites that mediate virus transmission were predicted through a network analysis. These predictions were confirmed through a micro-simulation that simulated virus transmission based on the actual contact history. The micro-simulations indicated that the majority of virus copies were transmitted through single items. This study contributes to the understanding of droplet and contact transmission in elementary schools and provides insights into infection prevention.

## Methods

### Data collection/ethics statement and privacy

The methodology used in this study was approved by the Institutional Ethics Review Board of the University of Tsukuba and Lion Corporation. In preparation for this survey, approval for the study from the cooperating school was obtained from the principal on 22-10-2022. The recruitment period for this study began on 30-11-2022 and ended on 22-12-2022. An information letter explaining the study was sent to the guardians of the participating students by the school principal at 30-11-2022. Written consent was subsequently obtained from the guardians of the participants. From 30-11-2022 to 08-12-2022, students and their guardians were allowed to opt out of the study. Furthermore, during the study period, until its conclusion on 22-12-2022, students and their guardians could withdraw their consent at any time. If consent was not given or was withdrawn, the respective guardians and students were instructed to submit a designated form, included in the information letter, to the teacher and the video recording team. The authors received only a report on the number of participants who opted out or withdrew consent.

The survey was conducted in a class at an elementary school in Tokyo, Japan, for four days in 12-2022. The students were 9–10 years of age and the class comprised 30 students. Communication and contact behaviors were videotaped using four and three cameras set in the classroom and hallway, respectively. The videos recorded the time from arrival to departure from the elementary school. The break time, excluding class hours (i.e., before morning and afternoon homeroom, three 5-minute breaks, one 10-minute break, and a lunch break) was used for the analyses. For privacy protection purposes, communication and contact behaviors were annotated from the recorded video data by a third-party institution. In addition, the video was pixelated after annotation by the third-party institution to prevent the identification of individuals. Each student was assigned an ID and analyzed anonymously once the data arrived at the researchers.

Communication behavior was annotated with the timestamp of the communication start, the ID of the student who initiated communications (the “initiator”), the student who was engaged in communication by the initiator (“target”), and the duration of the communication. Communication has be categorized into three types: conversation, contact (physical contact), and conversation and contact (both conversation and physical contact).

The annotation of the contact behavior included the student who contacted something; the contacted items including body parts (referred to as the “item”); and the owner of the item (either a personal belonging of a student or a shared common item). When a student contacted an item, it was recorded whether the individual was alone or in the presence of other people. These states were labeled “solo behavior” and “group behavior.” The layouts of the classroom and hallway are shown in Fig 1. Each faucet and hand wash were annotated individually and the items located at the front, back and sides of the classroom were also indicated individually.

**Fig 1.**
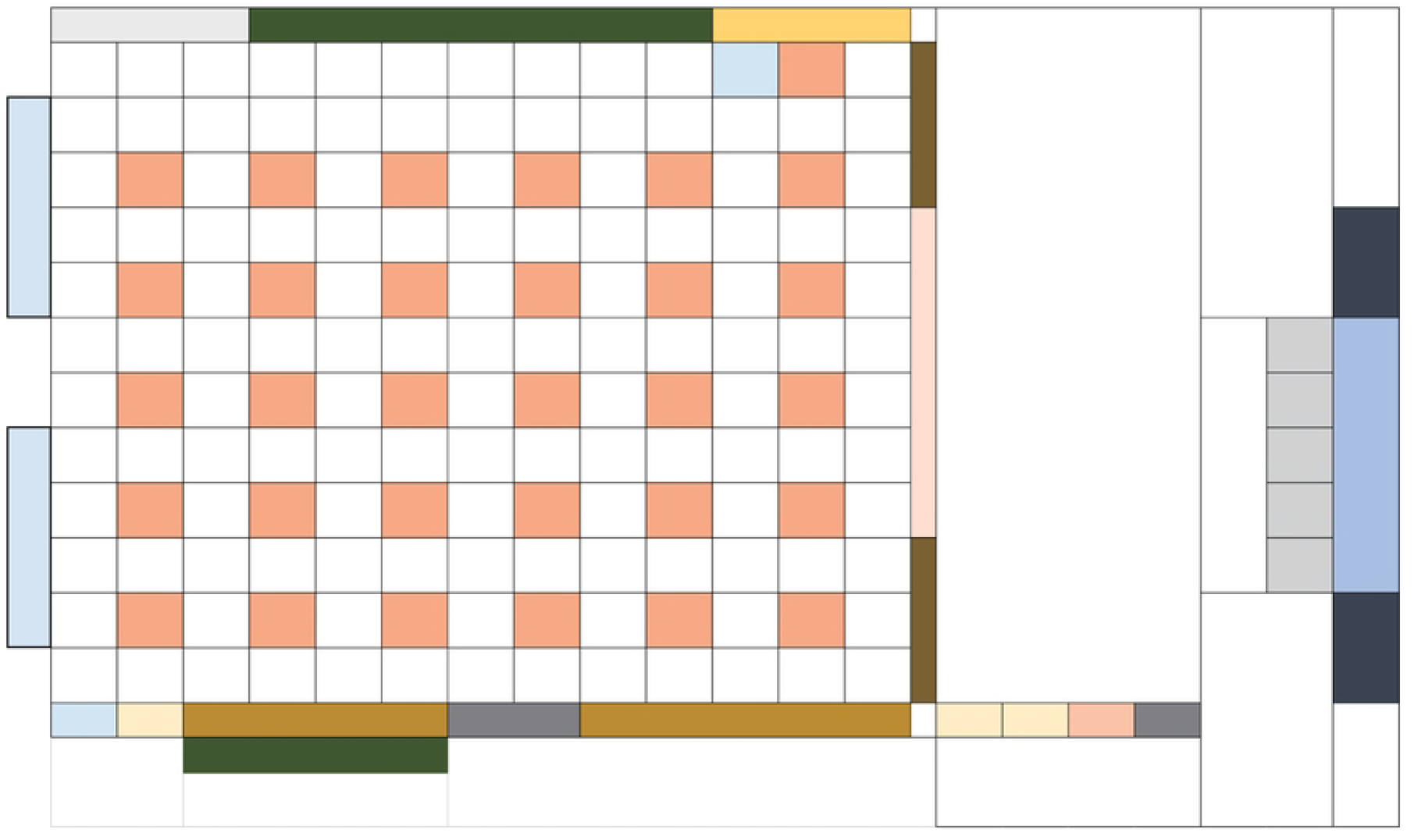
Layout of measurement area.

It should be noted that not all breaks during which the videos were recorded have been annotated.

### Network analyses

To analyze the communication and contact patterns among students, a network for each was generated.

The communication network was generated from the adjacency matrix of the initiators and targets. An initiator refers to a student who initiates communication, whereas a target refers to a student engaged in communication with the initiator. The communication network is represented as a directed graph, in which communication is conducted from the initiator to the target with a specific directionality. The degree of the communication network is the number of students who communicate during an arbitrary period. This value was counted if at least one communication was conducted between students (ID:*i*) and (ID:*j*). The in-degree and out-degree represent the number of initiators and targets from a student, respectively. The number of communications between a pair of students, (ID:*i*) and (ID:*j*), was counted during an arbitrary period.

The contact network was represented as an undirected graph. When a student (ID:*i*) contacted an item (ID:*x*), an edge was generated between the student’s hand (ID:*i*) and the item (ID:*x*). An example of a network is shown in Fig 2. The students’ hands and items are represented by black and orange circles, respectively. The degree is the number of students who contacted an item during an arbitrary period. The shortest distance refers to the shortest distance between the nodes of the student’s hand, computed using Dijkstra’s algorithm [24]. The shortest path betweenness centrality of node *c*_*b*_(*v*_*i*_) was calculated as follows [25]:

**Fig 2.**
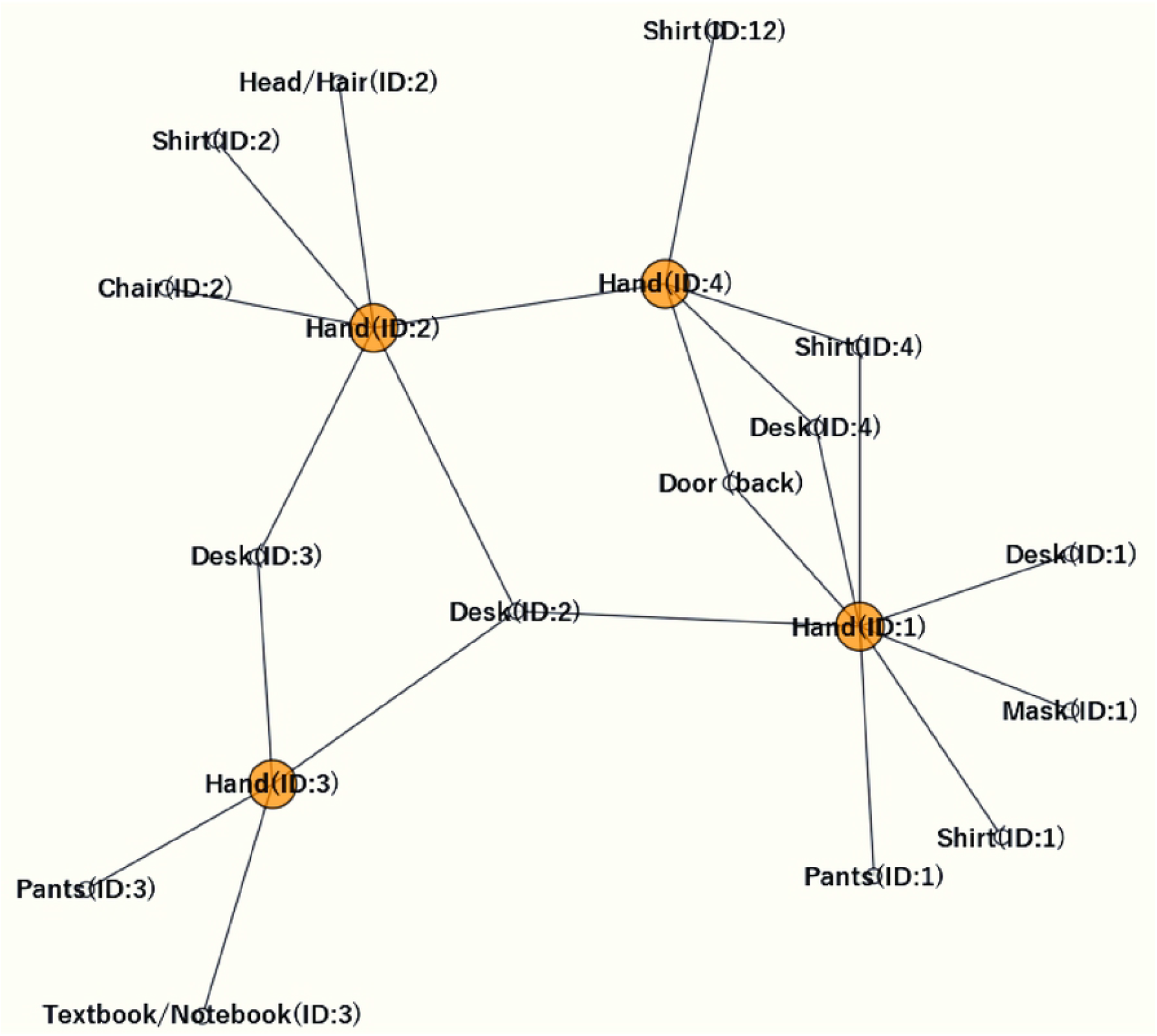
Contact network example. Orange and black circles represent students’ hands and items, respectively.

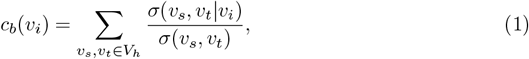

where *V*_*h*_ is the set of nodes of the student’s hand, *v*_*i*_, *v*_*s*_ and *v*_*t*_ are the *i*-th, start, and target nodes of the student’s hand, respectively. Moreover, *σ*(*v*_*s*_, *v*_*t*_) is the number of shortest paths between *v*_*s*_ and *v*_*t*_, *σ*(*v*_*s*_, *v*_*t*_|*v*_*i*_) is the number of those paths that pass through *v*_*i*_ other than *v*_*s*_ and *v*_*t*_.

The degree distribution indicates the characteristics of network. The probability *P* (*k*) at degree *k* is given by the following equation:

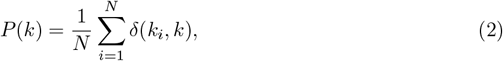

where *N* is the number of nodes, *k*_*i*_ is the degree of the *i*-th node. Furthermore, *δ*(*k*_*i*_, *k*) represent Kronecker delta function, which equals one when *k* = *k*_*i*_ and zero otherwise.

To visualize these networks, the Fruchterman–Reingold force-directed algorithm [26] was employed to determine the positions of the nodes. These network analyses and visualizations were conducted using the `NetworkX` library [27].

### Probability of infection through conversation

The probability of droplet infection of susceptible students through communication with virus-carrying students was calculated using an equation derived from numerical simulations [28–30].

The probability of droplet infection *P* is given by [29, 31, 32]:

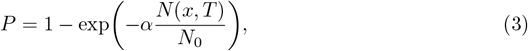

where *α* is a factor that regulates the infectivity caused by viral strains. *N*_0_ is assumed to be the average number of virus particles required to infect an individual. In this study, *N*_0_ was set to values ranging from 300 to 2000, with a focus on 900. This value has been used as *N*_0_ of SARS-CoV-2 in previous studies, and falls within a range similar to that of influenza A [29, 30, 33]. The number of inhaled virons, denoted as *N* (*x, T*), depends on the communication duration *T* and the distance *x* between the students. The number of inhaled virons, *N* (*x, T*) is given by

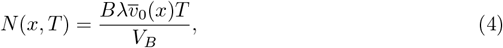

where *B* and λ are the breathing rate (*m*^3^/hr) of susceptible student and the viral load or viral density (copies/*m*^3^), expressed as the number of viral copies per unit volume of sputum. *V*_*B*_, was designated as a rectangular box with its long edge oriented in the direction from the nose of the susceptible student toward the ground. The probability of infection was evaluated by tracking droplets in their inhalation zones. The injection droplet volume that enters the inhalation zone *V*_*B*_ was the average inhalation rate of sputum droplets, represented by the initial droplet volume at distance *x*. To analyze the probability of infection for the students, the communication duration *T* was set as the total communication duration between pairs of students of breaks in a day.

### Micro-simulation

To analyze virus transmission through contact from virus-carrying students in elementary schools, a micro-simulation was conducted assuming the presence of viruses in the actual contact history.

Break time was selected as the contact behavior. Two cases of virus reduction were identified in the contact history of the students: handwashing using water in the handwashing area and disinfection using a disinfectant. In the micro-simulation, it was ensured that the number of virus copies on students’ hands would be reduced by 1*/*10^2^ and 1*/*10^4^ when washing hands with water and when disinfecting them, respectively [34, 35].

The virus transmission probability between contact materials and viruses and the approximate expression for the decreasing effect of multiple contacts were determined using values reported in previous studies [36, 37]. The items and materials used in this study are listed in S1 Table. The probability of virus transmission was calculated using the mean values from previous studies. These values were obtained through *in vitro* experiments using the influenza virus as a proxy for envelope viruses and a skin model made of protein leather, along with pieces of stainless steel, polypropylene, pottery, wood (veneered board), cardboard, wallpaper, cotton cloth, and model skin. Let *n* denote the number of contacts for a student, the number of virus copies adhering to the student’s hand, and the contact item, denoted as 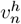 and 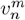, respectively, vary according to the following equations:

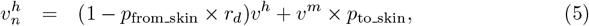

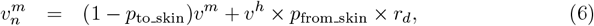

where *v*^*h*^ and *v*^*m*^, *p*_from skin_, *p*_to skin_ are the current number of virus copies adhering to the hand and item, the virus transmission probability from the hand to the material, and that from the material to the hand, respectively. The obtained values of 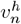 and 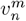 transform into *v*^*h*^ and *v*^*m*^ after the contact has ended. Additionally, *r*_*d*_ represents the rate of decrease and is given by

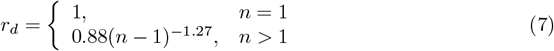

If the number of viruses adhering to the student’s hand and the item was less than one, it was considered as zero copies.

In a single simulation, one virus-carrying student was assigned, assuming that the virus present in the saliva of that student’s hand was deposited by coughing or sneezing immediately after the start of the break. Assuming a COVID-19 patient, it was hypothesized that 10^6^ copies of the virus would adhere to the hand [38].

## Results and Discussion

### Communication behavior

#### Communication behavior

The total number of communications and the communication duration during each break are presented in Table 1. The annotated communication behaviors are listed in S2 Table.

**Table 1.** Basic data of communication behavior.

| Break name (Date) | Measurement time | Total number of communication | Total communication duration (second) |
| --- | --- | --- | --- |
| Before morning homeroom (21-12-2022) | 8:04:51-8:15:46 | 152 | 2505 |
| 1st break (16-12-2022) | 9:22:05-9:26:25 | 31 | 513 |
| 2nd break (16-12-2022) | 10:09:40-10:34:00 | 119 | 2131 |
| 3rd break (20-12-2022) | 11:20:50-11:25:45 | 117 | 3288 |
| 3rd break (21-12-2022) | 11:22:15-11:25:29 | 27 | 379 |
| Lunch break (21-12-2022) | 12:04:27-12:23:57 | 259 | 4837 |
| Before afternoon homeroom (21-12-2022) | 14:01:11-14:13:04 | 51 | 613 |

The adjacency matrix of one day was generated using all the communication behaviors described in Table 1 (S1 Fig). In addition, a communication network generated from S1 Fig is shown in S2 Fig. The results were analyzed using data that incorporated the three types of communication. The adjacency matrix and network indicate the relationships among the students. These results show that the communication duration and number of communications varied across pairs of students. Furthermore, when a certain initiator communicated with a target, it did not necessarily engage in communication from the opposite perspective. To analyze the proportion of pairs of students engaged in bidirectional communication where they acted as both initiators and targets, the total degree of each student (which represents the degree of the undirected graph) and the degree of bidirectional communication are presented in Fig 3. The arithmetic average proportion of bidirectional communication among the 30 students was 0.498 with a standard deviation of 0.166. These results indicate that there is heterogeneity in the directionality of relationships based on communication among the students.

**Fig 3.**
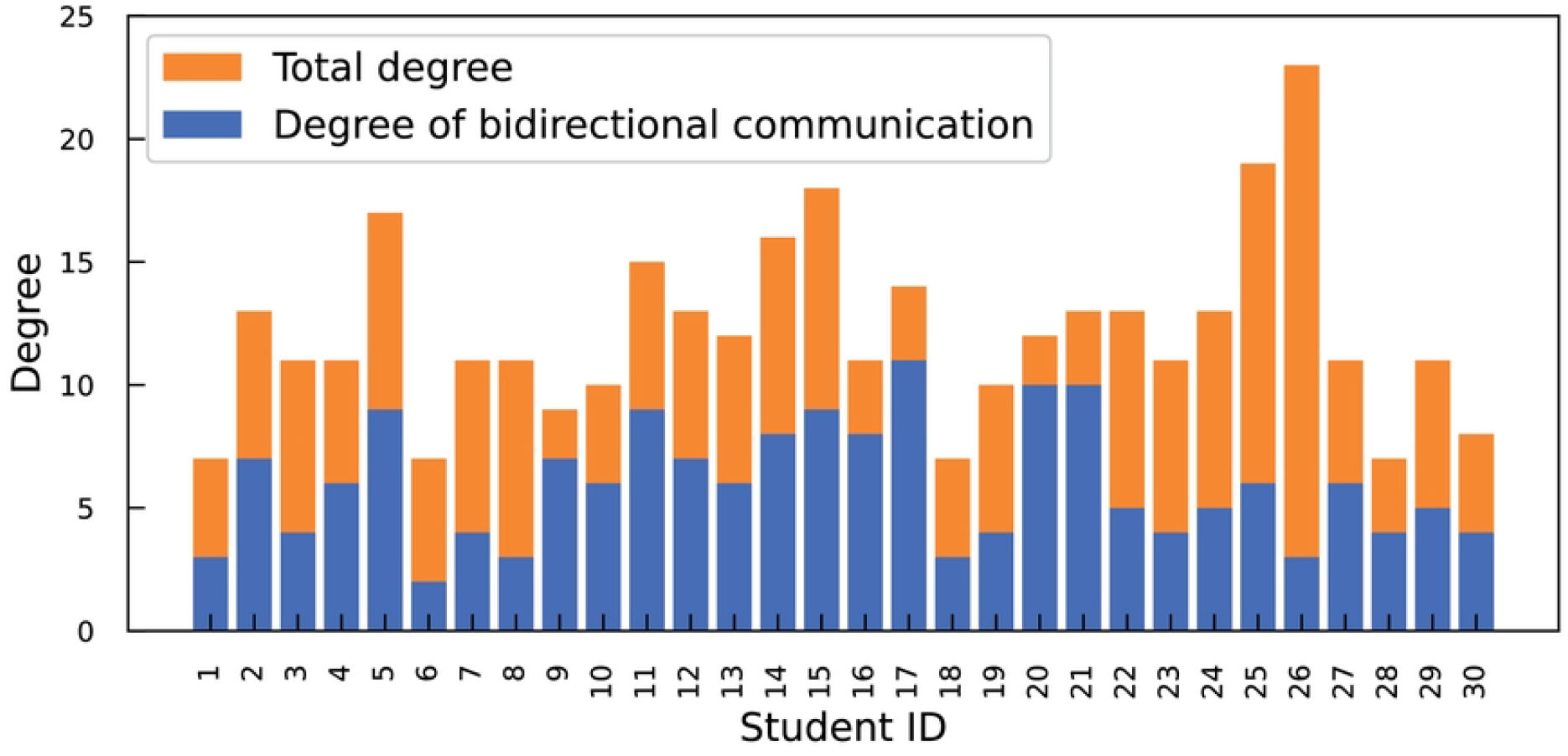
Proportion of pairs of students engaging in bidirectional communication.

The degree distributions were investigated to analyze the characteristics of the communication network. Figure 4 shows the degree distributions of both undirected and directed communication networks as well as the in-degree and out-degree of directed networks. The arithmetic average degrees of the original degree distribution, indicated by the black bar, were 12.1, 18.1, and 8.83 for the undirected, directed, and both in-degree and out-degree networks, respectively. In addition to the original degree distribution, a modified degree distribution was presented, wherein the degrees were grouped sequentially into sets of three starting from zero. The *P* (*k*) of the modified degree was the sum of the *P* (*k*) values for the three degrees. The representative value for the modified degree was set as the intermediate value among the three degrees. For example, when modifying the original degrees of 9, 10, and 11, the representative value of the modified degree was set to 10. The modified degree distribution suggests a distribution resembling a Poisson distribution, where the mode in the modified distribution is considered to be the mean value. These characteristics have also been observed in other elementary school communication network studies [21]. However, it was also found that the distribution of larger degrees would deviate from the Poisson distribution.

**Fig 4.**
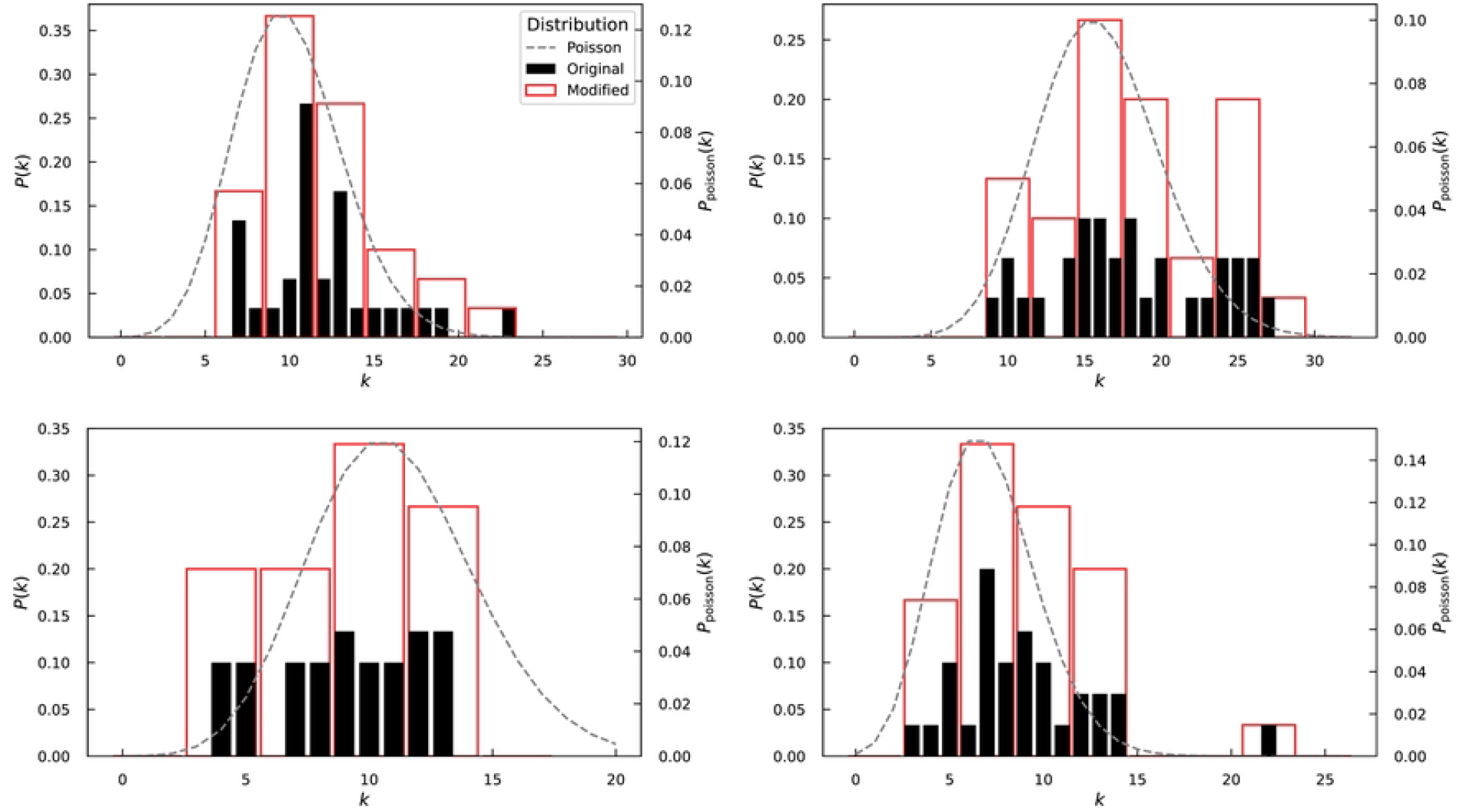
Degree distribution of communication networks. (a) Undirected network, (b) Directed network, (c) In-degree distribution of the directed network, (d) Out-degree distribution of the directed network. Black and red bars denote the original degree distribution and the degree distribution modified by grouping degrees into groups of three, respectively. The dashed line denotes the Poisson distribution where the mode in the modified degree distribution is considered as the mean value.

The distribution of communication duration and the number of communications are presented in Fig 5. Communication duration was analyzed based on values aggregated every 10 seconds. Representative values were assigned; for example, as 10 seconds for the range 1-10 seconds and 20 seconds for the range 11-20 seconds. To analyze only conversation time, the results of communication duration were used data excluding the “contact” communication type. Conversely, the results of the number of communications were analyzed using data that incorporated the three types of communication. For Fig 5, the distribution follows a power law, as observed in other elementary school communication network studies [19, 22]. These results indicate that most communications were of short duration and low frequency.

**Fig 5.**
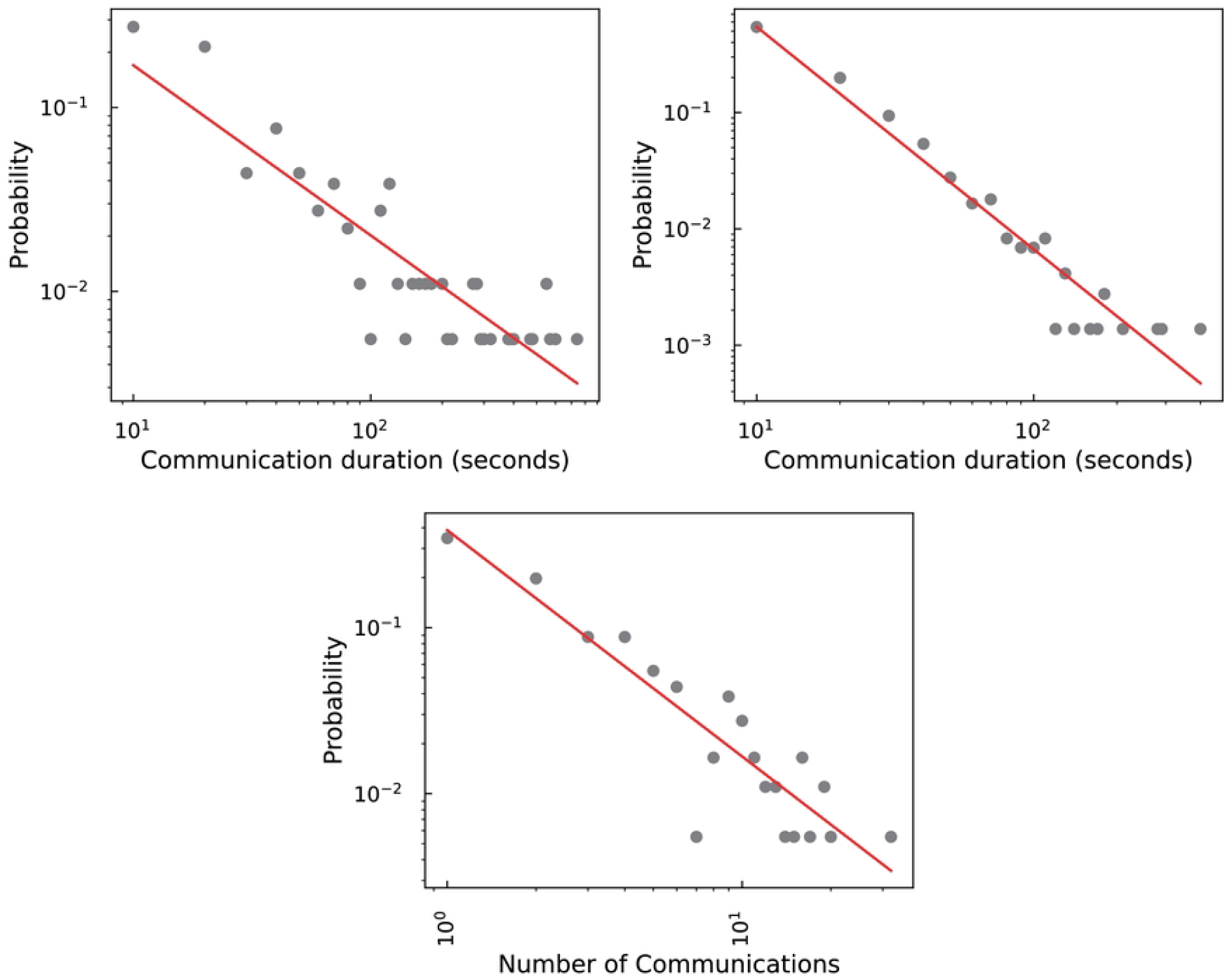
Distribution of communication duration and the number of communications of the total duration of breaks in a day. (a) Cumulative communication duration for each pairs of students, (b) communication duration of each conversation, (c) cumulative the number of communications for each pairs of students. The communication duration was analyzed by aggregating the values every 10 seconds. Red lines denote the approximating curves fitted with a power law.

#### Probability of infection through conversation

The probability of droplet infection through communication was calculated using an equation derived from numerical simulations [28–30]. When a student was a virus carrier, the total communication duration between the virus-carrying student and the susceptible student was used. As there were 30 patterns of infection probabilities from the virus-carrying students to the other 29 susceptible students, a total of 870 infection probabilities were calculated. Data on communication duration were used, excluding the “contact” communication type. The factor that regulated the infectivity caused by variant strains *α* was set to 1 or 5.77, as described in a previous study [30].

The arithmetic advantages of the probability of infection are listed in Table 2. It was suggested that increasing the distance between virus-carrying and susceptible students during conversations reduces the probability of infection. By quantifying the probability of susceptible students becoming infected when one of their classmates was a virus-carrying student, it was possible to assess the likelihood of transmission.

**Table 2.** Probability of infection through conversation when *N*_0_ = 900. The maximum and minimum probability correspond to *N*_0_ of 300 and 2000, respectively.

| Distance ($m$) | $P(\alpha = 1)(\%)$ | $P(\alpha = 5.77)(\%)$ |
| --- | --- | --- |
| 0.5 | 1.29<br>(0.754 – 3.10) | 5.05<br>(2.74 – 9.62) |
| 1.0 | 0.846<br>(0.568 – 1.85) | 3.18<br>(1.66 – 6.84) |

### Contact behavior

#### Characteristics of contact items

The cumulative number of contacts and number of contacted items during each break are shown in Table 3. The annotated contact behaviors are depicted in Table S3 Table. In the “Owner” column of S3 Table, the student ID was annotated when the contacted item belonged to a student. When the contacted item was a shared item, 0 is annotated.

**Table 3.** Basic data of contact behavior.

| Break name (Date) | Measurement time | Number of contacts |
| --- | --- | --- |
| Before morning homeroom (16-12-2022) | 8:04:15-8:18:27 | 1412 |
| 1st break (16-12-2022) | 9:20:21-9:25:55 | 567 |
| 2nd break (16-12-2022) | 10:09:36-10:43:23 | 1084 |
| 3rd break (20-12-2022) | 11:20:35-11:25:59 | 1032 |
| 3rd break (21-12-2022) | 11:20:52-11:25:59 | 604 |
| Lunch break (21-12-2022) | 12:07:51-13:19:58 | 4906 |
| Before afternoon homeroom (21-12-2022) | 14:01:40-14:09:48 | 371 |

The number of contacts for each item and item ownership in each break is presented in S4 Table. Excluding the “before afternoon homeroom,” desks had the highest number of contacts during all breaks. In the “before afternoon homeroom,” there was a higher frequency of contact with handbags, school bags, and outerwear as students prepared to go home. Similarly, those items were contacted during the “before morning homeroom” because of the period of arrival at elementary school. During the “lunch break,” there was a higher aggregated number of contacts (Table 3 “Lunch break”). This can be attributed to the longer measurement time and the fact that students collectively served lunch, leading to increased interaction among students. Additionally, items such as tableware, milk, and lunch mats were considered lunch specific. Later during the “lunch break,” as each student needed to wait until all classmates had finished eating individually, students came into contact with their tablets. Throughout multiple breaks, desks and shirts had a high frequency of contact not only with personal ownership by oneself (self) but also with belongings owned by others (others). As for the common items, there was a higher frequency of contact with the door, faucet, desk (teacher), and serving table (i.e., serving table cover) located at front of the classroom (Fig 1). The door was frequently touched during movement between the classroom and hallway, while the faucet was contacted for handwashing and drinking water in the handwashing area. The desk (teacher) and serving table had a higher frequency of contact because of the accumulation of students around the front area of the classroom, where the teacher was present. Here, since the “3rd break (20-12-2022)” had a higher number of contacts per unit of time, the top 20 contacted items are shown as representatives (Fig 6). These results suggest the potential transmission of viruses through contact with belongings owned by others or through shared items.

**Fig 6.**
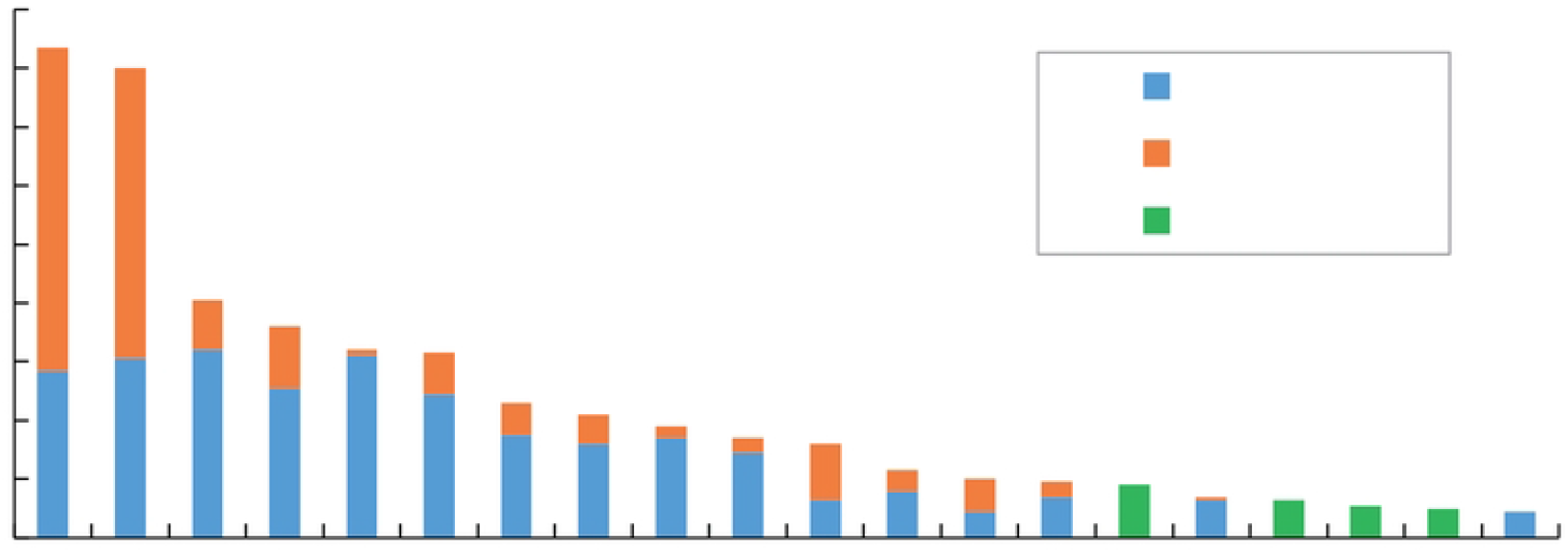
Top 20 contacted items at the “3rd break (20-12-2022).”

To elucidate the contact with items and belongings owned by others, the number of contacts with items and ownership of each item during group and solo activities were analyzed. The data during the group and solo activities during each break are presented in S5 Table and S6 Table, respectively. As a representative example, the results of the “3rd break (20-12-2022)” are portrayed in Fig 7. Common items were excluded from the analysis. According to these results, it was demonstrated that students were in contact with belongings owned by others during group activities and mostly in contact with their own items during solo activities. This suggests a potential impact of contact behavior on virus transmission during group activities.

**Fig 7.**
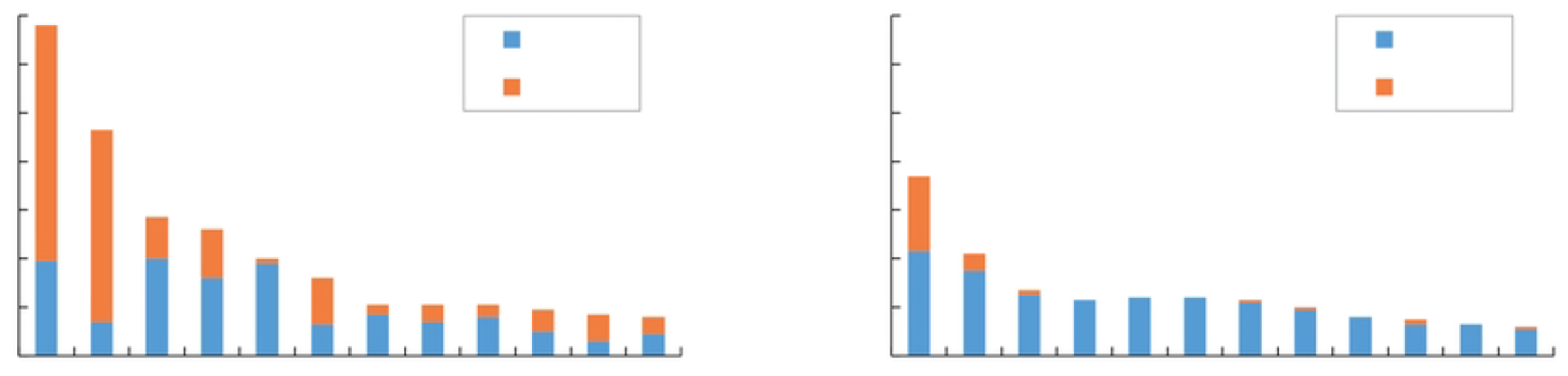
Contacted items the “3rd break (20-12-2022).” (a) Group activities, (b) solo activities. The items with the number of contacts *>* 10 are indicated.

#### Network analysis

To estimate the potential for virus transmission through contact behaviors, contact behaviors were represented using a network. The contact network created from the contact behaviors during the “3rd break (20-12-2022)” is presented in Fig 8.

**Fig 8.**
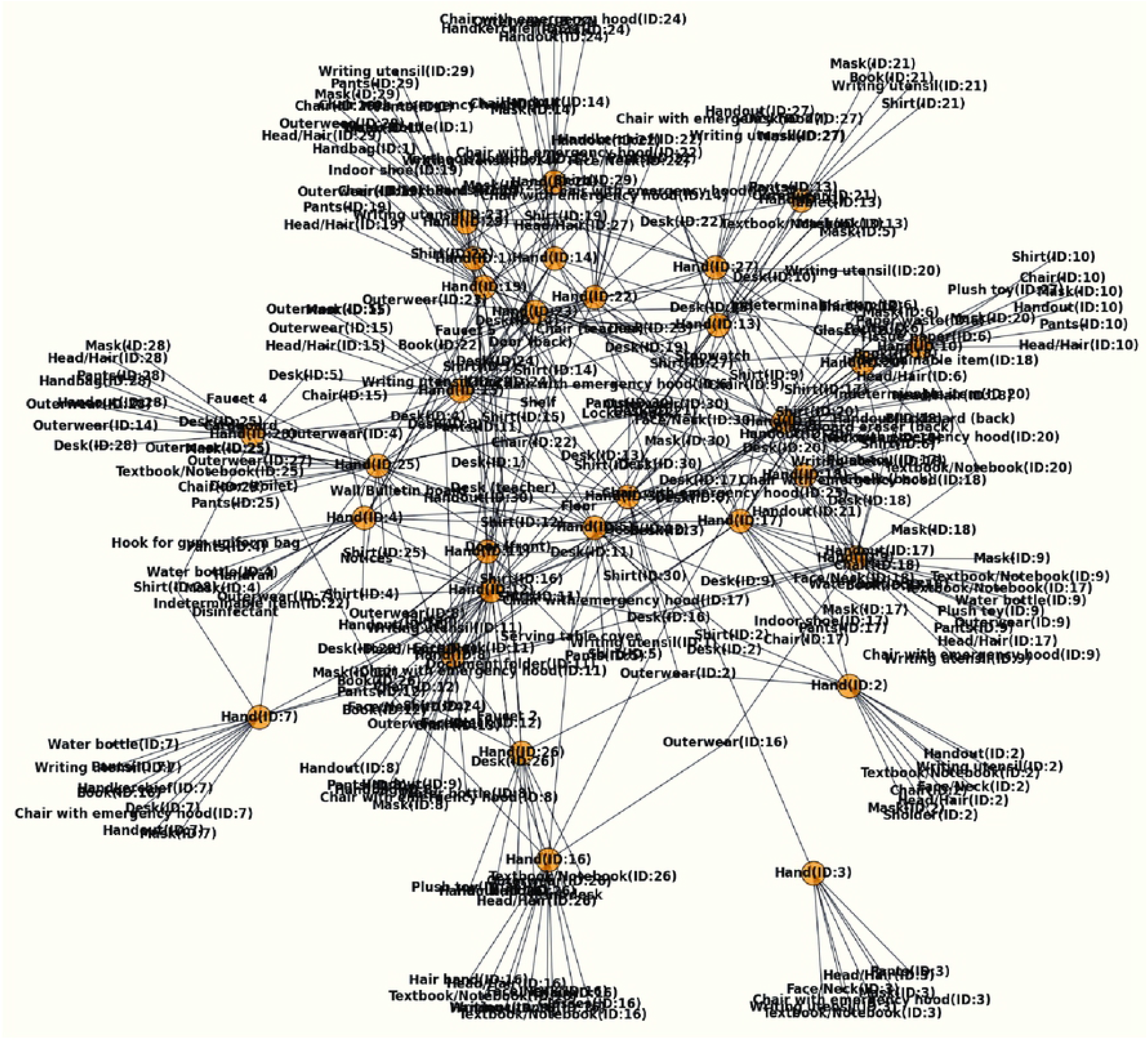
Network of contact behavior using “3rd break (20-12-2022).” Orange circles represent students’ hands.

First, the shortest path between students’ hands was investigated. The results are shown in Fig 9. The hand of the starting-point student was not included in the path. Therefore, when the path was 1, it represented the direct connection between the student’s hands, and when the path was 2, it indicated the connection through one item. In the contact network, the percentages of Paths 1, 2, 3, 4, and 5 were 2.07%, 50.8%, Path 3 was 15.4%, Path 4 was 31.5%, and Path 5 was 0.230%, respectively. Path 2 had the highest occurrence, indicating the potential for virus transmission through a single item in most situations. This suggests that the transmission between student’s hands may not require a large number of items.

**Fig 9.**
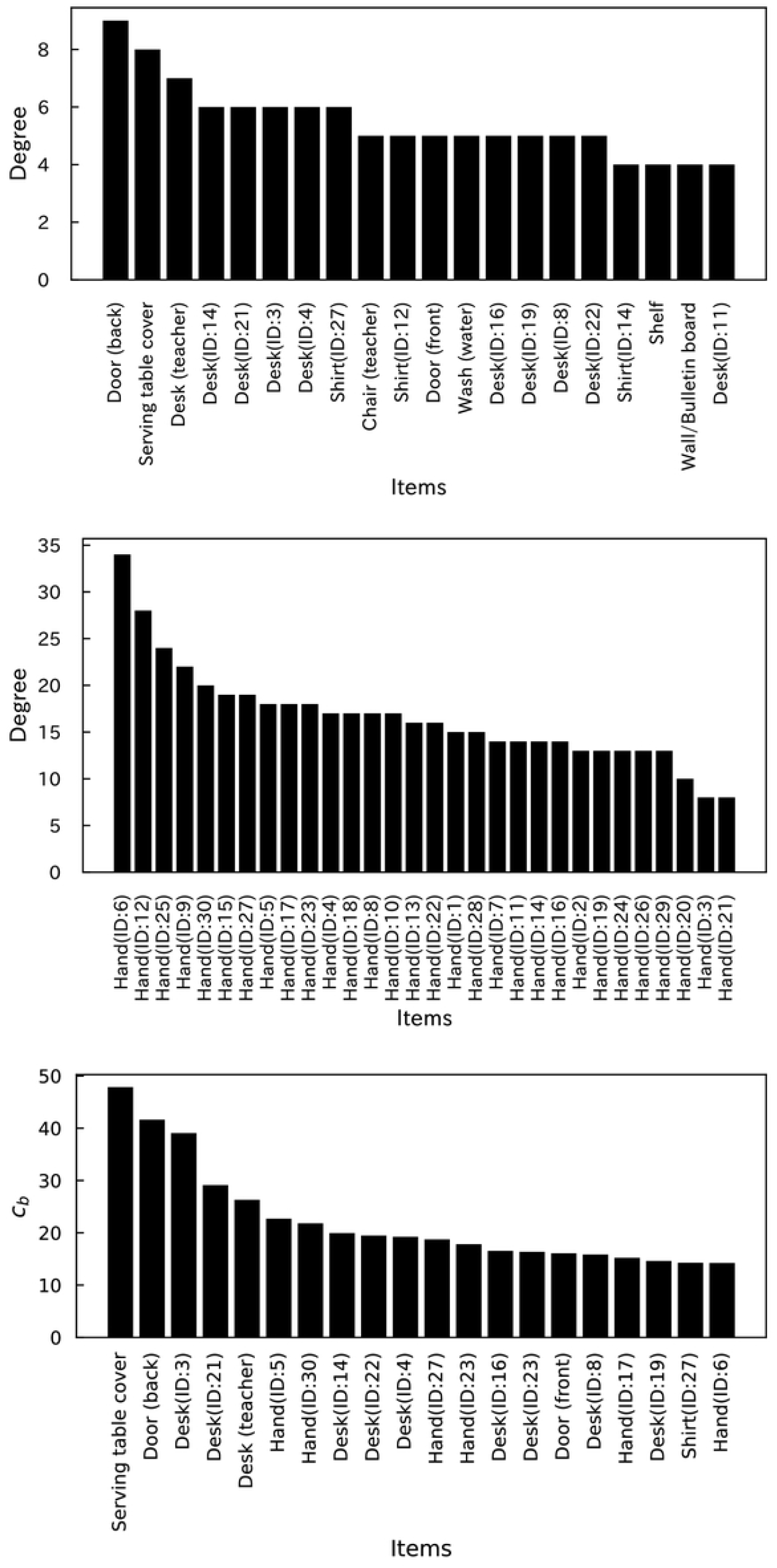
Heatmap of shortest path between student’s hands. The numerical values written within the heatmap represent the shortest path between students’ hands. The hand of the starting-point student is not included in the count of the numbers.

Next, to predict the candidate items that can be fomites of virus transmission, network metrics such as degree and betweenness centrality were investigated. The top 20 items of the value of degree, excluding students’ hands and betweenness centrality, and the degree of only the students’ hand in the network are shown in Fig 10. The total values in Fig 10 are presented in S7 Table. Because the order of the hands and other items was different, a degree analysis was conducted separately. Incidentally, the degree distribution of the items excluding hands exhibited a power-law-like distribution (S3 Fig). According to Fig 10, common items such as the serving table cover, desk (teacher), and door (back) had higher degrees and betweenness centrality. Personal belonging items, such as desks, shirts, and hands, exhibited higher degrees and betweenness centrality. These items were contacted at a higher frequency, as described in the previous subsection (Fig 6). These results suggest that these items have the potential to be used as fomites for virus transmission.

**Fig 10.**
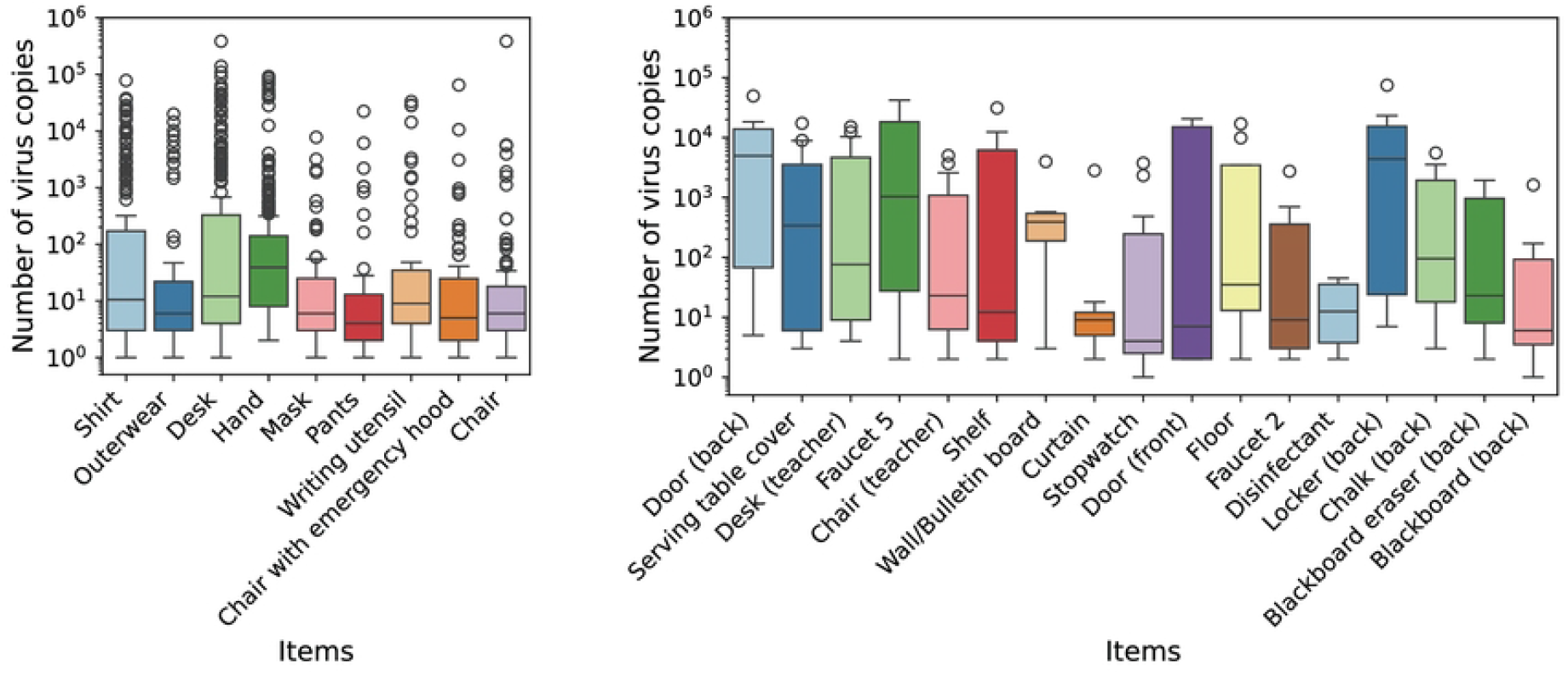
Network metrics of network of contact behavior. (a) Degree of top 20 items without students’ hands, (b) degree of only students’ hands, (c) betweenness centrality of top 20 items.

#### Analysis of contact infection with micro-simulation

In the previous subsection, potential fomite candidates for virus transmission through contact were predicted based on the contact network. Therefore, in this subsection, we assume the presence of the virus in the actual contact history and analyze the plausibility of the previous subsection using a micro-simulation.

A micro-simulation was conducted, assuming one student as a virus carrier, and it was performed for 30 individuals. The contact history of “3rd break (20-12-2022)” was selected as the contact behavior because all students’ hands were connected through fomites in this history (Fig 9). In each simulation, the number of virus copies transmitted to items other than their own belongings (including hands) was recorded, and the results are displayed using a box plot (Fig 11). Only the results where the virus adhered to items with ≥ 1 copies were included in the figure. The results for the belonging of others and common items are presented separately. The number of samples where ≥ 1 copies were adhered to each item is presented in S8 Table. Fig 11 displays the items that were sampled more than 50 for belongings of others and more than five for common items. In the case of belonging of others, because the maximum number of samples was 870 (30 times 29), the number of samples with virus adherence was higher than that of the common items. From Fig 11(a), when the virus adhered to the belongings of others, the median number of virus copies for each item was approximately 10^1^. For desks, shirts, and hands, which were identified as potential fomites in the previous subsection, the 75th percentile value exceeded 10^2^ copies. This suggests that these components were more susceptible to virus adherence. Furthermore, the maximum number of virus copies adhering to these items was greater than that of other items. Moving on to the common items, although the sample size was smaller, the median number of virus copies surpassed that of the belongings of others. Items such as doors, serving table covers, and desks (teachers), which were identified as potential fomites in the previous subsection, had larger sample sizes and higher 75th percentile (Fig 11(b)).

**Fig 11.**
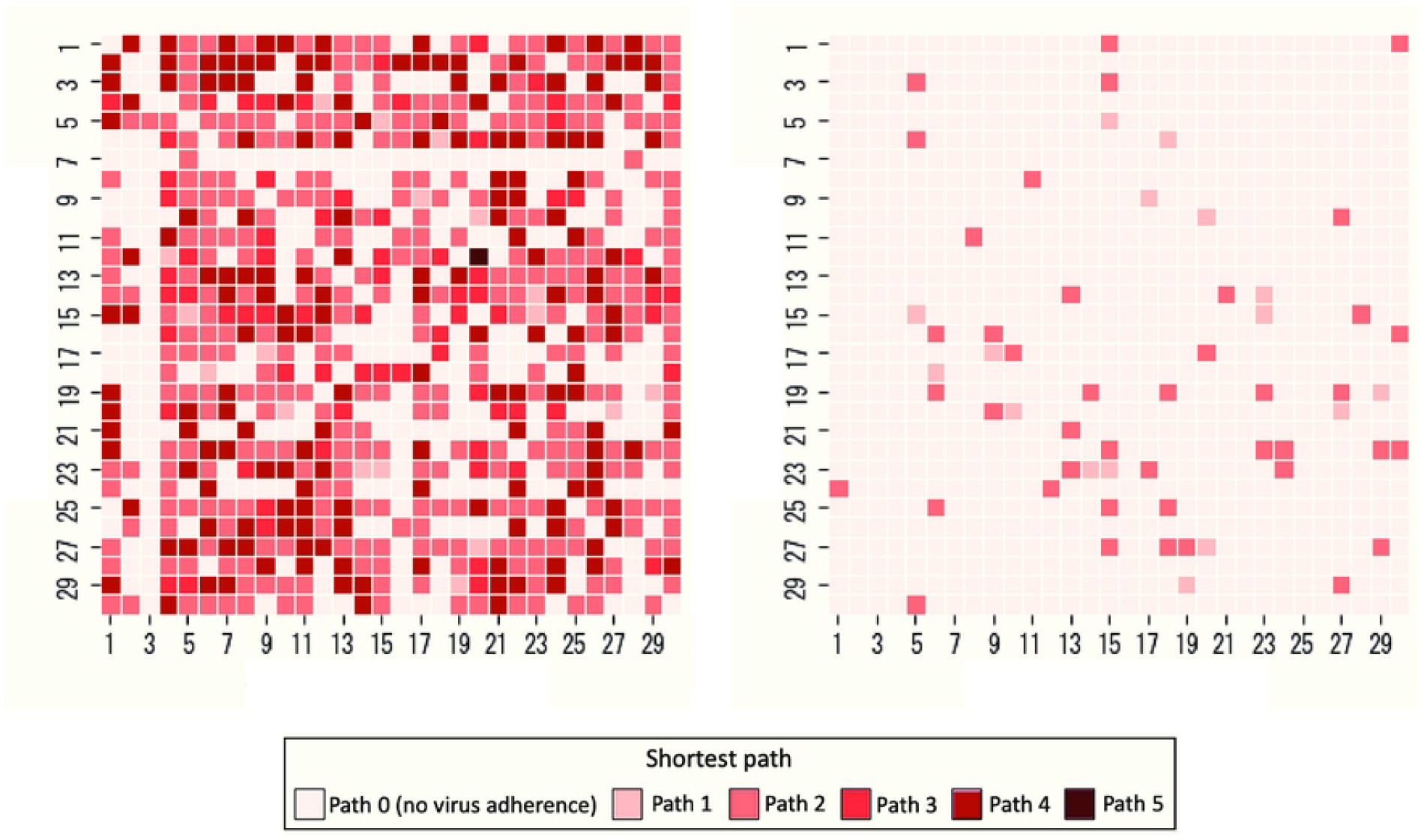
Relationship between items and adhered virus copies. (a) Items belongings of others, (b) common items.

Next, to analyze the relationship between virus transmission and infection, the students’ hands were examined. Fig 12 illustrates the susceptible students who had the virus adhered to their hands by a virus-carrying student at the end of the simulation. The figure indicates susceptible students with virus adherence of *≥* 1 copies, as well as those with virus adherence of *≥* 300 copies. The value of *≥* 300 copies represents the minimum average number of virus particles *N*_0_. Additionally, for each virus-carrying a student simulation, a contact network was formed based on the items that had virus adherence through contact (referred to as the “virus transmission contact network” and shown in S4 Fig), and the shortest paths between the hands of the students was measured.

**Fig 12.**
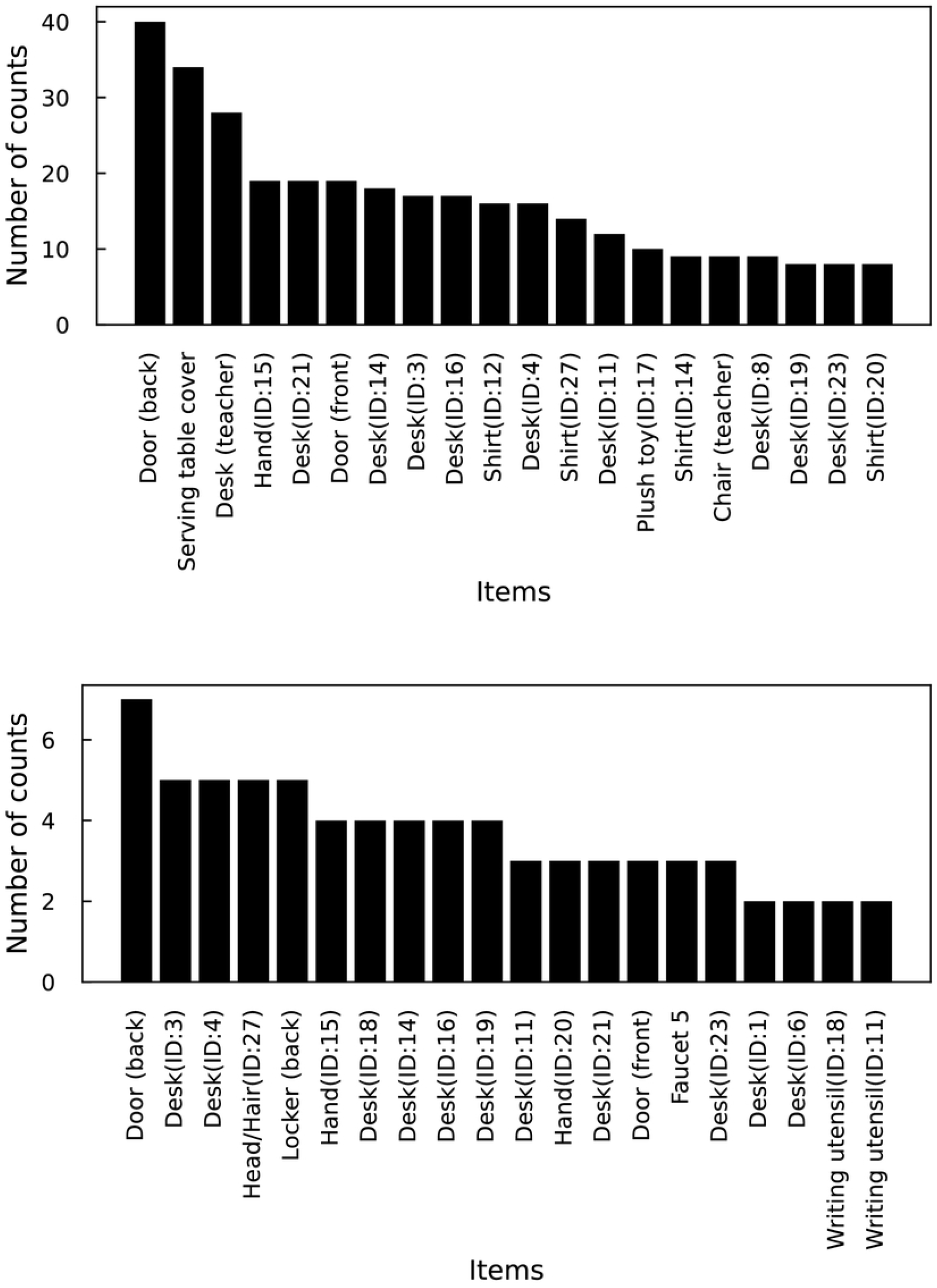
Heatmap of the susceptible students who had the virus adhered to their hands by a virus-carrying student at the final of the simulation. Susceptible students with virus attachment of (a) ≥ 1 and (b) ≥ 300.

The arithmetic average number of susceptible students with virus adherence of ≥ 1 copies was 19.8 (Fig 12(a)). The maximum was 26 and the minimum was two. Except when ID 5 was a virus carrier, the susceptible hand of ID 3 did not adhere to the virus. This result is consistent with the finding that ID 3 had the highest arithmetic average shortest path for each student (Fig 9). The percentages of the shortest path among all susceptible students with virus adherence were investigated. The percentages of Paths 1, 2, 3, 4, and 5 were 3.04%, 54.6%, Path 3 was 12.1%, Path 4 was 30.0%, and Path 5 was 0.169% (Fig 12(a)). Compared to the distribution of the shortest path in Fig 9, which indicated the potential for virus transmission, the distribution of the virus transmission contact network showed a higher proportion of Paths 1 and 2. This suggests that a more efficient transmission pathway is the path through a smaller number of items. The arithmetic average number of susceptible students with virus adherence of ≥ 300 copies was 1.93 (Fig 12(b)). The maximum and minimum were five and zero, respectively. In the virus transmission contact network, the shortest path was of only two types: Path 1 was 27.6% and Path 2 was 72.4% (Fig 12(b)). It was found that the transmission of a high number of virus copies predominantly occurred through a single item. This is attributed to the decay in the number of virus copies being transmitted gradually due to the decreasing rate associated with contact frequencies and low virus transmission probability. It has been demonstrated that in contact-based virus transmission, the presence of a single item in contact with the hands of a virus-carrying student, where a significant amount of the virus has adhered, is crucial.

Finally, the items that transmitted the virus in the micro-simulation were analyzed. The items that transmitted ≥ 1 or ≥ 100 copies of the virus to the hands after contact and the number of transmissions by the items are shown in Fig 13. The top 20 fomites are also indicated in the figure. The total relationship between the fomites and the number of transmissions is listed in S9 Table. It should be noted that when the pairs of virus-carrying students, susceptible students, and items were the same, they were not counted. For instance, if the virus-carrying student ID: *i* adhered the virus to a door and the susceptible student ID: *j* contacted the door multiple times, resulting in multiple instances of virus transmission, the number of transmissions by the fomite in that case was considered to be one. Additionally, the reason for selecting the threshold of ≥ 100 for virus transmission is that it was not possible to explain all cases where ≥ 300 copies of the virus were adhered to the hands of other student using a higher threshold.

**Fig 13.**
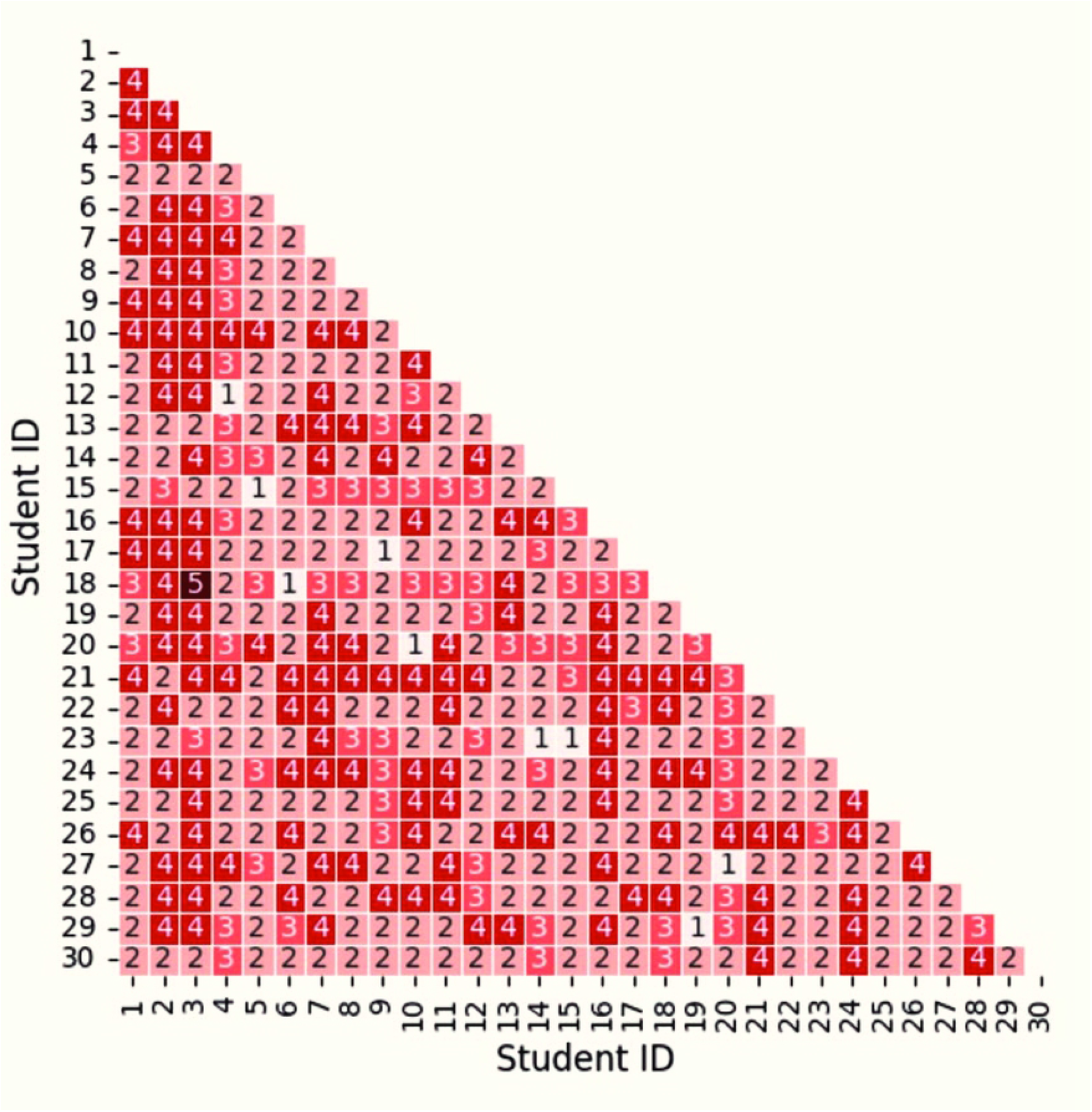
Fomites and the number of transmission in micro-simulation. The fomites that transmitted (a) ≥ 1 or (b) ≥ 100 copies of the virus to the hands after contact. Top 20 fomites are shown.

In fomites where ≥ 1 copies were transmitted, the door, serving table cover, and desk (teacher) had a higher number of counts. This aligns with the results of the analysis of the contact network’s degree and betweenness centrality (Fig 10). Among fomites where ≥ 100 copies were transmitted, the door had a higher number of counts, while the serving table cover and desk (teacher) did not transmit the virus (S9 Table). This can be attributed to the material of the items. The door, serving table cover, and desk (teacher) were made of stainless steel, cotton cloth, and wood (veneered board), respectively, with average virus transmission probabilities of 0.49%, 0.11%, and 0.20%, respectively. Considering the low virus transmission probabilities of items other than the doors, it can be concluded that a higher number of virus copies were not transmitted. Furthermore, although the locker (back) has a higher number of counts, it could not necessarily be considered an important item because it was contacted as a single item despite its wide range of contact. In terms of personal belonging, it appeared that items such as desks, shirts, and hands, which had a high frequency of contact and were associated with high network metrics, could serve as potential fomites.

## Limitations

In this study, the communication and contact behaviors were annotated by humans using video recordings. The accuracy of the data may be reduced in situations where the video quality was poor or where students were clustered closely together. Because most students wore masks, the presence or absence of conversations was determined based on the situation; therefore, the reliability of the data can be considered limited.

This study was conducted in one class in a single elementary school. Thus, it is possible that the results are specific to this particular environment. Additionally, this study captured behaviors in winter, during which time the government had issued recommendations regarding COVID-19. Since the COVID-19 pandemic had been ongoing in Japan for approximately three years since its onset in 2020, it is believed that students had a heightened awareness of infection prevention. Therefore, it might not reflect the natural and näıve behavior of elementary school students. Nevertheless, there are consistencies with prior studies, but these results should be treated with caution.

The contact area is not considered in the micro-simulation. Both virus-carrying and susceptible students contact the same area of each item. In reality, the contact area would not be limited; therefore, it is important to note that the results of virus transmission might be based on excessive conditions.

## Conclusion

This study investigated communication and contact behaviors in an elementary school and analyzed their relevance to droplet and contact transmission. The analysis of communication behaviors revealed the heterogeneous nature of communication among students. The risk of droplet transmission in elementary schools was quantified by calculating droplet transmission probabilities based on conversation duration. In the analysis of contact behavior, a novel approach was established to create networks based on contact history. This enabled the prediction of items with the potential to serve as fomites for viral transmission, as demonstrated by the analysis of contact networks. The reliability of these predictions was further supported by micro-simulations. Furthermore, the results of the micro-simulations indicated that the majority of the viral copies were transmitted through single items and did not spread beyond that. In particular, the analysis of contact transmission is unprecedented, making this research valuable in providing effective information on infectious disease prevention in elementary schools.

In previous studies, the risk of virus transmission was investigated using simulations, and an agent-based model was employed to simulate the behavior of one or two individuals within a household and proposed appropriate infection control measures [36, 37]. This study examined the behaviors of elementary school students in relation to infection risk. These insights will contribute to the construction of simulation models for analyzing infection risks in elementary schools. We plan to construct models and explore suitable infection control measures in elementary schools.

In some previous studies, the risk of virus transmission using simulations has been conducted. We constructed an agent-based model to simulate the behavior of one or two individuals within households and proposed the appropriate infection control measures [36, 37]. This study examined the behavior of elementary school students in relation to infection risks. The insights contribute to the construction of simulation models for analyzing infection risks in elementary schools. We will construct the models and explore suitable infection control measures at elementary schools.

## Supporting information

**S1 Fig. Heatmaps of the adjacency matrix from communication behavior of breaks in a day**. (a) Communication duration, (b) the number of communication of pair between initiators and targets.

**S2 Fig. Networks from communication behavior of the total duration of breaks in a day**. (a) Communication duration, (b) the number of communication of pair between initiators and targets. The arrows denote the direction of communication from the initiator to the target. The thickness of the arrows in (a) and (b) represents the abundance of communication duration and the number of communications, respectively.

**S3 Fig. Distribution of contacted items excluding hands**. Red and blue lines denote the approximating curves fitted with a power law.

**S4 Fig. Example of virus transmission contact network**. The red and orange circles represent the virus-carrying students’ hand and the susceptible students’ hands with virus adherence of ≥ 1. The shortest paths between the hands of virus carriers and susceptible students were measured. For instance, the shortest path between ID:28 and ID:7 was Path 2.

**S1 Table. Items and materials**.

**S2 Table. Communication behaviors. S3 Table. Contact behaviors**.

**S4 Table. The number of contact for each item and item ownership in each break**.

**S5 Table. The number of contact during the group activities in each break. S6 Table. The number of contact during the solo activities in each break. S7 Table. Network metrics of network of contact behavior**.

**S8 Table. The number of samples with *≥* 1 copies**.

**S9 Table. The relationship between the fomites and the number of transmissions**.

## Data Availability

All relevant data are within the manuscript and its Supporting Information files.

## Acknowledgments

We express our gratitude to the participants, guardians, and teachers who participated in this survey. We would also like to extend our gratitude to Professor Tsubokura and Dr. Rahul from Kobe University and RIKEN for generously providing us with the droplet infection probability function.

